# A prospective study to identify rates of SARS-CoV-2 virus in the peritoneum and lower genital tract of patients having surgery

**DOI:** 10.1101/2020.07.30.20165043

**Authors:** Dominique Jones, David Faluyi, Sarah Hamilton, Nicholas Stylianides, Ken Ma, Sarah Duff, Nicholas Machin, Richard J Edmondson

**Affiliations:** Manchester University NHS Foundation Trust, Oxford Road, Manchester, UK; Division of Cancer Sciences, Faculty of Biology, Medicine and Health, University of Manchester, Saint Mary’s Hospital, Manchester, UK

**Keywords:** SARS-CoV-2, laparoscopy, peritoneal fluid, gynaecology, colorectal surgery

## Abstract

**Introduction:** The risks to surgeons of carrying out aerosol generating procedures during the COVID pandemic are unknown. To start to define these risks, in a systematic manner, we investigated the presence of SARS-CoV-2 virus in the abdominal fluid and lower genital tract of patients undergoing surgery.

**Methods:** We carried out a prospective cross sectional observational study of 113 patients undergoing abdominal surgery or instrumentation of the lower genital tract. We took COVID swabs from the peritoneal cavity and from the vagina from all eligible patients. Results were stratified by pre operative COVID status.

**Results:** In patients who were presumed COVID negative at the time of surgery SARS-CoV-2 virus RNA was detected in 0/102 peritoneal samples and 0/98 vaginal samples. Peritoneal and vaginal swabs were also negative in one patient who had a positive nasopharyngeal swab immediately prior to surgery.

**Conclusions:** The presence of SARS-CoV-2 RNA in the abdominal fluid or lower genital tract of presumed negative patients is nil or extremely low. These data will inform surgeons of the risks of restarting laparoscopic surgery at a time when COVID19 is endemic in the population.

## Introduction

As health services around the world emerge from the first wave of COVID 19 infections the ability to resume normal work patterns becomes of paramount importance. Surgeons in particular need to be able to deliver effective and safe treatments to patients and reduce the inevitable backlogs that have developed.

Laparoscopy and other surgical procedures such as colposcopy and hysteroscopy are essential parts of the management of cancer patients with laparoscopy being associated with reduced hospital stay and reduced complication rates compared to open surgery [1].

However the safety of these procedures, which are known to be aerosol generating, is unclear, thus leading to a profusion of conflicting guidance (reviewed in [2]). Although the presence of other viruses, including Hepatitis B virus, has been confirmed in laparoscopic aerosols [3] evidence for the presence of SARS-CoV-2 virus is mixed [4] and has not been studied systematically.

The first step in such a process is to determine the risk of undertaking minimal access surgery in a low risk patient. Here we describe a prospective study to determine the prevalence of SARS-CoV-2 virus in peritoneal fluid, a prerequisite for the presence of viral particles in laparoscopic aerosol or surgical plume, and the lower genital tract.

## Methods

### Study population

Patients were enrolled into the study at Saint Mary’s Hospital, Manchester Royal Infirmary and Wythenshawe Hospital, all part of Manchester University Foundation NHS Trust, between 7^th^ June and 29^th^ July, 2020. The study was registered with the National Institute of Health Research (NIHR) register of COVID related trials. Ethical approval was obtained from Frenchay Research Ethics Committee (20/SW/0088) and informed written consent obtained from all patients prior to the collection of swabs and data. The study was sponsored by Manchester University NHS Foundation Trust.

### Inclusion and exclusion criteria

Patients were eligible if they were undergoing abdominal surgery including laparoscopy, laparotomy and caesarean section, or undergoing instrumentation of the lower genital tract including but not limited to colposcopy, hysteroscopy and insertion of intra uterine device. All patients had a nasopharyngeal COVID swab taken within 48 hours of the procedure.

Patients were excluded if there was evidence of faecal or amniotic contamination of the peritoneal cavity prior to the swab being taken

### Study procedures

Where applicable patients had one or both of the following samples collected.

A vaginal COVID swab was taken from all four vaginal fornices, prior to any vaginal prep, catheterisation or vaginal instrumentation

An abdominal COVID swab was taken as soon as the abdominal cavity was opened. The swab was taken from the pelvis and the paracolic gutters. For laparoscopic cases, the swab was inserted through a 5mm operating port, manipulated using laparoscopic forceps to ensure it was applied firmly to peritoneal surfaces and then removed immediately through the laparoscopic port.

Samples were collected in viral transport medium and tested for the presence of SARS-CoV-2 RNA by reverse-transcriptase polymerase chain reaction (RT-PCR) using the Cobas® SARS-CoV-2 RNA assay (Roche, USA).

### Clinical Risk Groups

For the purposes of analysis patients were classified into the three risk groups defined by NHS England [5]. Many UK centres have adopted these risk groupings as a mechanism for cohorting patients within hospitals. For this study the three groups were therefore defined as:

Green (COVID-clear). Patients who self isolate for 14 days prior to admission, undergo negative nasopharyngeal swab within 48 hours of admission (and in the case of patients being admitted to level 2 and 3 care also undergo negative chest imaging) and are treated by teams of clinicians only caring for “green” patients.

Amber (undifferentiated). Patients who have not self isolated but are asymptomatic at time of admission, undergo nasopharyngeal testing within 4 hours of admission or have suspicious but not diagnostic chest radiological findings.

Blue (COVID-positive). Patients who have had positive COVID nasopharyngeal testing, or who have symptoms or radiological findings in keeping with current acute infection.

### Clinical COVID testing

All patients recruited underwent standard screening for active COVID-19 infection using combined nose and throat swabs, tested for SARS-CoV-2 RNA by RT-PCR.

Whilst the study was recruiting SARS-CoV-2 antibody testing became available for some patients and these results, where available, were collected from the patient record along with other clinical details.

### Population infection rates estimation

Estimates of the incidence of COVID positivity during the study were derived from UK government websites.

### Sample size and statistical analysis

As this was an observational study a formal power calculation was not possible. The number to be recruited was therefore estimated using a Bayesian approach. The number needed for the study was defined as the number needed to convince a majority of clinicians that the rate of COVID positivity in a particular cohort is sufficiently low that the benefits of surgery outweigh the risks.

Polling a panel of surgeons revealed that a rate of 0/100 for any particular risk group would be sufficiently low risk to convince surgeons that this was a safe procedure. A total number of 100 cases was therefore selected for the study.

Data were analysed using MS Excel.

Data are reported in line with STROBE guidance for the reporting of cross sectional observational studies [6].

## Results

A total of 113 patients were recruited, of whom 102 and 98 had a peritoneal and/or vaginal swab taken respectively. Only 1/113 patients had a positive nasopharyngeal swab prior to surgery. This patient was asymptomatic at the time of admission for elective caesarean section, underwent the procedure as planned, and has remained well in the post operative period.

### Peritoneal swab cohort

Peritoneal sampling was carried out in 102 patients at the time of surgery. Clinical characteristics of this cohort are shown in table 1. Of note, during the study period there was only one patient undergoing surgery who had evidence of acute infection (blue risk). 78 were managed as amber risk and 23 as green risk. 4/31 (13%) of this cohort who underwent testing showed evidence of SARS-CoV-2 antibody suggesting previous infection.

**Table 1:** patient characteristics (peritoneal cohort)

|  |  |  |
| --- | --- | --- |
| Total cohort |  | 102 |
| Age (median & range) |  | 36 (20-83) |
| ethnicity | White<br>BAME<sup>1</sup> | 70<br>32 |
| BMI<sup>2</sup> | <25<br>25-40<br>>40<br>Not known | 31<br>59<br>1<br>11 |
| Operative procedure | Caesarean section<br>Cancer surgery<br>Other | 72<br>19<br>11 |
| Antibody testing | Not tested<br>negative<br>positive | 71<br>27<br>4 |
| COVID cohort<sup>3</sup> | Green<br>Amber<br>Blue | 23<br>78<br>1 |

1 BAME- black, asian and minority ethnic
2 BMI- body mass index
3 COVID cohort as defined by NHS England (see text [5])

0/65 peritoneal swabs were positive for SARS-CoV-2 virus RNA. This included the one patient who had a positive nasopharyngeal swab immediately prior to caesarean section.

### Vaginal swab cohort

Vaginal sampling was carried out in 98 patients at the time of surgery. Clinical characteristics of this cohort are shown in table 2. Of note, during the study period there was only one patient undergoing surgery who had evidence of acute infection (blue risk). 75 were managed as amber risk and 22 as green risk. 4/32 (13%) of this cohort who underwent testing showed evidence of SARS-CoV-2 antibody suggesting previous infection.

**Table 2:**
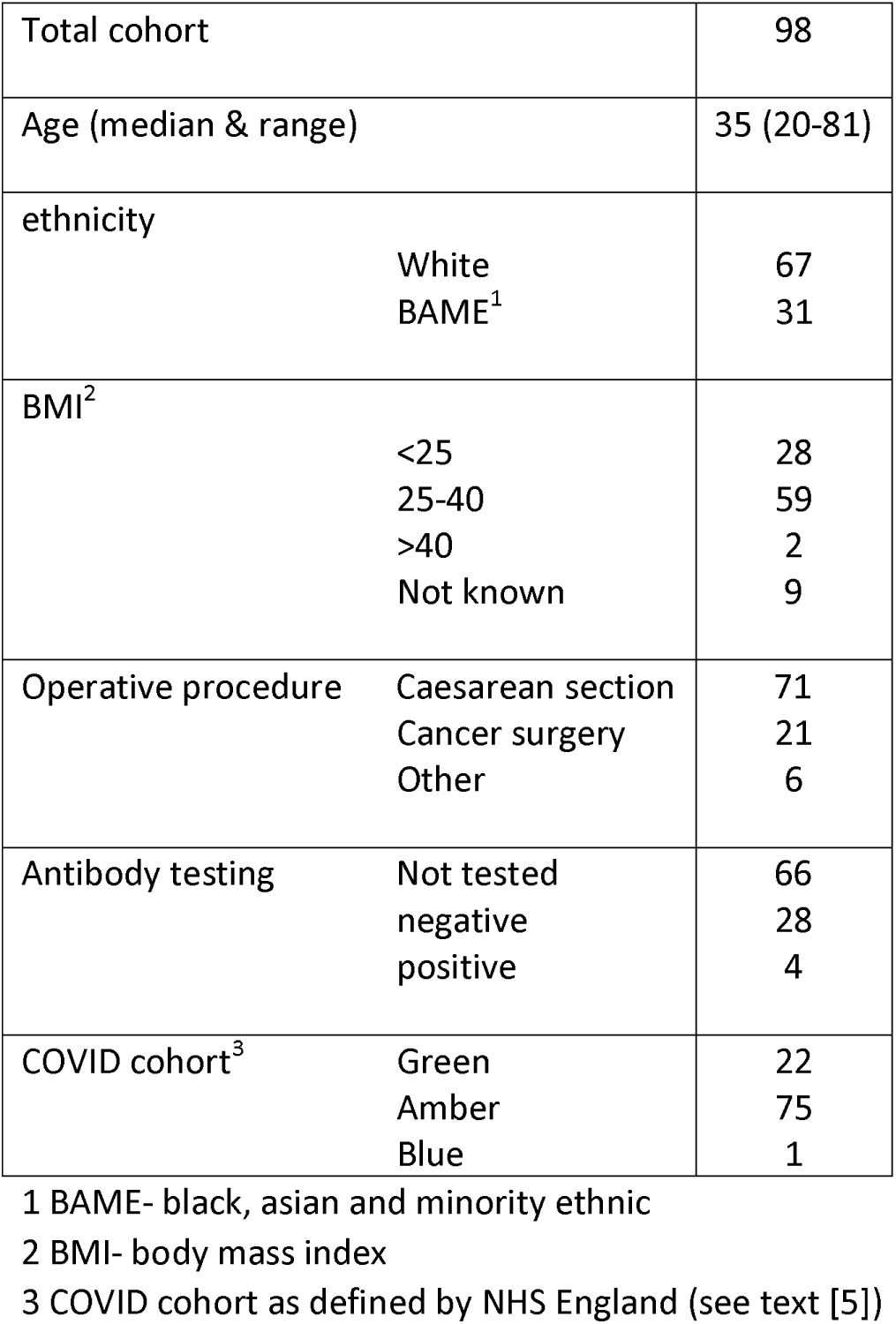
patient characteristics (vaginal cohort)

0/98 vaginal swabs were positive for SARS-CoV-2 virus RNA. This included the one patient who had a positive nasopharyngeal swab immediately prior to caesarean section.

## Discussion

In this study we were unable to detect any SARS-CoV-2 RNA in either the abdominal cavity or the vaginal fluids of a prospective series of over 100 patients undergoing surgery.

Specifically, the numbers of “green” (n=27) and “amber” (n=84) patients included here provides reassurance that the risks in these two groups are extremely low and that surgery in patients who have been tested preoperatively could be undertaken safely, including minimal access surgery as appropriate.

It is also interesting that we were unable to detect virus in the 4/32 (13%) patients who had positive antibody titres and the one patient who had active infection at the time of surgery. These numbers remain too small to make strong recommendations but we will continue to recruit these subgroups of patients for further study.

This study was carried out at a time when the community incidence rate of COVID infection was estimated to be stable at 0.02% [7]. This rate of infection would seem to represent the ongoing endemic rate which is likely to persist for at least a period of months.

Our study was weighted towards the inclusion of patients undergoing caesarean section as this is currently the commonest abdominal operation being carried out in our institution. There is no reason to suppose that this population is particularly different from the general surgical population.

This study has not included patients in whom there was faecal contamination of the abdominal cavity at the time of surgery. Further studies are now underway to address this and also to assess risk in asymptomatic and symptomatic COVID positive patients, and those who have positive antibodies.

In summary we have demonstrated that undertaking abdominal and gynaecological surgery, including laparoscopic surgery, in patients who have been COVID tested pre operatively, whether they have been self isolating or not, must be considered a low risk procedure. These data will allow surgeons, and their teams, to make informed decisions about returning to laparoscopic surgery, the role of filtration devices, and the need for PPE in low risk patients who comprise the majority of patients currently awaiting a surgical procedure.

## Data Availability

no non patient identifiable data are available from this study

## Declarations

None of the authors have any conflicts of interest to declare

